# DNA methylation derived immune cell profiles, CpG markers of inflammation, and pancreatic cancer risk

**DOI:** 10.1101/2020.01.27.20019034

**Authors:** Dominique S. Michaud, Mengyuan Ruan, Devin C. Koestler, Lola Alonso, Esther Molina-Montes, Dong Pei, Carmen J. Marsit, Immaculata De Vivo, Núria Malats, Karl T. Kelsey

## Abstract

Pancreatic cancer is projected to become the second most common cause of cancer death over the next 5 years. Since inflammation is thought to be a common trajectory for disease initiation, we sought to prospectively characterize immune profiles using DNA methylation markers to examine whether they play a key role in pancreatic cancer risk. In a nested case-control study pooling three U.S. prospective cohort studies, DNA methylation was measured in prediagnostic leukocytes of incident pancreatic cancer cases and matched controls using the Illumina MethylationEPIC array. Differentially methylated regions were used to predict immune cell types and CpGs previously associated with blood inflammatory markers were selected for the analysis. DNA methylation data from a retrospective case-control study conducted in Spain (PanGenEU) was used for independent replication of results. Immune cell proportions and ratio of cell proportions were not associated with pancreatic cancer risk in the nested case-control study. Methylation extent of CpGs residing in or near gene *MNDA* was significantly associated with pancreatic cancer risk in the nested case-control study and replicated in PanGenEU. In the nested case-control study, the associations were present 10 or more years prior to cancer diagnosis. Methylation of a promoter CpG of gene *PIM-1* was associated with pancreatic cancer survival in both studies. We identified several CpGs that may play a role in pancreatic carcinogenesis using a targeted approach for the selection of inflammation-based CpGs in two large, independent studies conducted in different countries with distinct study designs.

## Introduction

In the absence of specific disease symptoms, pancreatic cancer is difficult to identify early in the course of the disease; only 10% of pancreatic tumors are localized at diagnosis.^1^ Overall mortality for pancreatic cancer is very high, with only 9% of patients surviving 5-years beyond diagnosis, primarily because over 50% of cases have metastasized by diagnosis,^1^ making tumors inoperable. Identifying pancreatic cancer at earlier stages could significantly improve survival with increased opportunities for surgery; however, due to poor diagnostic accuracy of existing detection methods, screening is currently not recommended for asymptomatic adults.^2^

New high-dimensional arrays designed to measure DNA methylation levels at hundreds of thousands of CpG sites throughout the genome have opened opportunities to estimate immune cell proportions in frozen blood samples that were stored without the measurement of complete blood counts (CBC) or without assessing immune profiles.^3^ With this method, archived samples from prospective studies can be used to examine changes in the immune response in individuals who develop cancer months or years later, providing new opportunities to better understand biological mechanisms and, perhaps, identify biomarkers for early detection.

Immune cell proportions, such as the ratio of neutrophil to lymphocyte (NLR), have been shown to accurately predict cancer survival,^4, 5^ but no study has evaluated whether immune response markers based on DNA methylation profiles are associated with risk of developing pancreatic cancer. To address this, we examined associations between known DNA methylation markers of immune response and pancreatic cancer risk using pre-diagnostic bloods of cases and controls obtained from three large US cohort studies. Results were then replicated in a large Spanish case-control study; replication in a completely different study population provides an opportunity to evaluate generalizability of our findings, rule out any bias, or chance findings that may have occurred in the discovery nested case-control study.

## Methods

The analysis described in this paper represents two different study designs: a nested case-control dataset sampled from 3 U.S prospective cohort studies, and a retrospective case-control study conducted in Spain (PanGenEU). The primary analyses were conducted on pancreatic cancer cases and matched controls identified from the Nurses’ Health Study (NHS), the Physician’s Health Study (PHS), and the Health Professionals Follow-up Study (HPFS). The secondary analyses, intended to replicate the findings from the nested case-control study, were conducted using pancreatic cancer cases and controls from Spanish participants of the PanGenEU study, a multicenter case-control study based in Europe. More details for each study are provided in the **Supplementary Methods**.

In the cohort studies, 403 incident cases were confirmed to have pancreatic cancer among the participants who provided blood samples prior to cancer diagnosis. A control subject was matched to each case on cohort (which also matches on sex), age (+/-1 year), date of blood draw (month 3+/- and year), smoking (never, past, current) and race (White/other). Incident density sampling was used for the selection of controls. A subset of participants had data on inflammatory markers (C-reactive protein, interleukin-6 and tumor necrosis factor-alpha) from a prior study in the same cohorts.^6^ The final dataset consisted of 393 cases and 431 controls. For the survival analysis, cases missing date of diagnosis (n=42) or date of death (n=9) were not included in the analysis.

The replication dataset was conducted using pancreatic cancer cases and controls obtained from the Spanish component of the European Study into Digestive Illnesses and Genetics (PanGenEU), a multicenter case-control study that was conducted between 2009-2014 in six European countries (Spain, Italy, Germany, United Kingdom, Sweden and Ireland).^7-10^ For the methylation analyses, we selected a PanGenEU representative subset of 657 Spanish subjects, 357 cases and 300 controls. The final data set for this analysis included a total of 338 cases and 285 controls.

### DNA methylation measurements

DNA extracted from buffy coats (nested case-control study) or leukocytes (PanGenEU) were bisulfite-treated and DNA methylation was measured with the Illumina Infinium MethylationEPIC BeadChip array (Illumina, Inc, CA, USA). Details on DNA methylation measurements and data processing are provided in the **Supplementary Methods**. Reproducibility of results from 850K Illumina array has been previously shown to be very high (r=0.997).^11^ In addition, we previously conducted a pilot study to examine reproducibility of DNA methylation measured in peripheral blood over a 1-year period using this array and demonstrated that DNA methylation varies by site, but is stable across a large number of probes.^12^

### Estimation of immune cell composition

Leukocyte subtypes proportions (i.e., CD4T, CD8T, natural killer cells [NK], B cells, monocytes [Mono] and neutrophils) were estimated using the “*estimateCellCounts2*” function in the FlowSorted.Blood.EPIC Bioconductor package,^13^ which is based on previously published reference-based cell mixture deconvolution algorithm with reference library selection conducted using the IDOL methodology.^14^

### Inflammation-associated CpG sites

We selected 64 CpG sites that had been strongly associated with inflammation markers in previous studies to examine in this study.^15, 16^ Eleven CpGs from Ahsan et al^15^ were associated with multiple inflammatory blood markers among 698 individuals (listed in their Table 1), and 54 CpG sites reaching EWAS significance (4 CpG sites were not on the 850K array) from a large EWAS conducted to identify DNA methylation markers for C-reactive protein levels.^16^ Of those, 1 CpG overlapped with the other publication. Finally, we removed 14 CpGs that had low ICCs in our pilot study. ^12^ The remaining 50 CpGs we tested had ICCs ranging between 0.40 and 0.95 (calculated from the M values adjusted for age, cell composition and Combat adjusted). The CpGs with significant associations (in our results) had ICCs between 0.67 and 0.86.

**Table 1.** Baseline characteristics for study population, by study and case-control status at end of follow-up

| Mean (SD) or N (%) | Total cohort studies (N=824) |  | NHS (N=370) |  | HPFS (N=297) |  | PHS (N=157) |  | PanGenEU (N=623) |  |
| --- | --- | --- | --- | --- | --- | --- | --- | --- | --- | --- |
|  | Cases | Controls | Cases | Controls | Cases | Controls | Cases | Controls | Cases | Controls |
| **N** | 393 | 431 | 176 | 194 | 146 | 151 | 71 | 86 | 338 | 285 |
| **Age at study entry** | 60.6 (7.9) | 60.2 (7.7) | 59.2 (6.4) | 59.5 (6.2) | 63.8 (7.9) | 63.6 (7.8) | 57.5 (9.2) | 55.8 (8.3) | 66.3 (12.7) | 63.7 (13.2) |
| **Time before diagnosis (year)<sup>a</sup>** | 13.0 (6.2) |  | 14.3 (6.3) |  | 10.9 (5.6) |  | 13.8 (6.3) |  | N/A |  |
| **Female** | 176 (44.8%) | 194 (45.0%) | all female |  | all male |  | all male |  | 142 (42.0) | 127 (44.6) |
| **Race<sup>b</sup>** |  |  |  |  |  |  |  |  |  |  |
| White | 346 (94.0%) | 406 (94.9%) | 164 (93.2%) | 187 (96.4%) | 137 (93.8%) | 145 (96.0%) | 45 (97.8%) | 74 (89.2%) | 334 (98.8) | 276 (96.8) |
| Black | 4 (1.1%) | 1 (0.2%) | 1 (0.6%) | 1 (0.5%) | 2 (1.4%) | 0 | 1 (2.2%) | 0 | 1 (0.29) | 2 (0.70) |
| Other | 18 (4.6%) | 21 (4.9%) | 11 (6.3%) | 6 (3.1%) | 7 (4.8%) | 6 (4.0%) | 0 (0.0%) | 9 (10.5%) |  |  |
| **Smoking<sup>c</sup>** |  |  |  |  |  |  |  |  |  |  |
| Never | 159 (40.9%) | 173 (40.3%) | 73 (42.0%) | 81 (41.5%) | 55 (38.2%) | 58 (39.2%) | 31 (43.7%) | 34 (39.5%) | 137 (40.5) | 141 (49.5) |
| Past | 174 (44.7%) | 190 (44.3%) | 73 (42.0%) | 81 (41.5%) | 77 (53.5%) | 76 (51.4%) | 24 (33.8%) | 33 (38.4%) | 97 (28.7) | 80 (28.1) |
| Current | 57 (14.6%) | 65 (15.2%) | 29 (16.6%) | 32 (16.5%) | 12 (8.3%) | 14 (9.5%) | 16 (22.5%) | 19 (22.1%) | 100 (29.6) | 61 (21.4) |
| **BMI (kg/m<sup>2</sup>)<sup>d</sup>** | 26.0 (4.3) | 25.6 (3.9) | 26.2 (5.3) | 25.8 (4.7) | 25.9 (3.2) | 25.9 (3.3) | 25.9 (3.1) | 24.7 (2.6) | 27.1 (4.6) | 27.12 (5.6) |
| **Diabetes<sup>e</sup>** | 19 (4.8%) | 11 (2.6%) | 11 (6.2%) | 7 (3.6%) | 4 (2.7%) | 2 (1.3%) | 4 (5.6%) | 2 (2.3%) | 108 (32.0) | 49 (17.2) |
| **hsCRP (mg/L)<sup>f</sup>** | 2.92 (5.53) | 2.83 (5.41) | 4.16 (7.48) | 3.48 (6.55) | 2.56 (4.30) | 2.46 (4.29) | 1.38 (1.24) | 2.25 (4.50) |  |  |
| **TNF-αR2 (pg/mL)<sup>g</sup>** | 2602 (677) | 2546 (645) | 2812 (691) | 2717 (640) | 2619 (665) | 2582 (698) | 2236 (505) | 2234 (455) |  |  |
| **IL-6 (pg/mL)<sup>h</sup>** | 2.2 (4.2) | 1.9 (3.3) | 2.7 (5.2) | 2.0 (4.8) | 1.8 (2.8) | 1.5 (1.0) | 1.9 (3.9) | 2.1 (2.5) |  |  |
| **Adiponectin (ng/mL)<sup>i</sup>** | 6674 (4759) | 6761 (4098) | 8716 (6080) | 8033 (4863) | 5158 (2464) | 6135 (3592) | 5380 (3248) | 5544 (2609) |  |  |

SD: standard deviation, BMI: body mass index, hsCRP: high sensitivity C-reactive protein, TNF- αR2: tumor necrosis factor-receptor, IL-6: interleukin-6.
<sup>a</sup> 11 missing values; <sup>b</sup> missing values for cohorts: 28, and PanGenEU: 10; <sup>c</sup> missing values for cohorts: 6, and PanGenEU: 7; <sup>d</sup> missing values for cohorts: 14, and PanGenEU: 44; <sup>e</sup> missing values for cohorts: 1, and PanGenEU: 4; <sup>f,g</sup> 336 missing values; <sup>h</sup> 349 missing values; <sup>i</sup> 341 missing values; no data for PanGenEU for serum biomarkers.

### Statistical analyses

All statistical analyses were performed in R (version 3.5.1). Immune cell ratios (e.g., CD4/CD8, neutrophil/lymphocyte, B cell/lymphocyte, T cell/lymphocyte) were calculated for each sample by taking the ratio of its predicted cell proportions described above. Quartiles were assigned according to distribution of immune cell ratios among controls. A series of unconditional multivariable logistic regression models were used to evaluate the association between immune cell ratio and pancreatic cancer case/control status (unconditional models were selected to maximize power by including controls without matched cases; results using conditional regression models were compared and no differences were observed for the ORs). Age at blood draw, cohort, smoking status (never, former, current), and date of blood draw (continuous) were adjusted for in each model. To minimize loss of cases/controls due to missing data, we did not include BMI as a covariate in the model; moreover, including BMI in sensitivity analyses did not alter associations. Similar models were used to examine the association between inflammation-associated CpG sites (modeled as quartiles; study specific) and pancreatic cancer case/control status. In addition to adjusting for previously mentioned covariates, these models were additionally adjusted for cell composition (e.g., estimated proportions of CD4T, CD8T, NK, B cell and monocytes) given the potential for confounding by cell composition.^17^

For the nested case-control study, Spearman’s rank correlation was used to calculate the correlation between methylation beta-values and C-reactive protein, IL-6 and TNF-alpha^6^ (Supplemental Table 1), as the biomarker and methylation beta-values were not always normally distributed. Correlations between methylation beta-values of inflammation CpG probes were also estimated using Spearman’s rank correlation (Supplemental Figure 2).

We examined the association between survival time and both immune cell ratios and the 50 inflammation CpGs among cases in the cohort studies using a series of multivariable Cox proportional hazard models. Age at blood draw, cohort, smoking status, date of blood draw, and time between blood draw and cancer diagnosis were adjusted for in the Cox proportional hazard models. Models testing for associations with inflammation-related CpG sites were additionally adjusted for estimated cell composition as described above. Associations with methylation levels were tested using tertiles and trends were tested using median values of each tertile entered as a continuous variable.

## Results

Characteristics of the participants included in this analysis are provided in Table1; due to matching criteria in the cohorts, age and smoking status were similar in cases and controls. On average, participants in the nested case-control study were diagnosed with pancreatic cancer at 60.6 years old, and provided blood samples an average of 13 years (range 6 months to 26 years) prior to diagnosis (Table 1 presents range for each study). Those who later developed pancreatic cancer had a slightly higher BMI than those who did not develop pancreatic cancer (BMI 26.0 vs 25.6 kg/m^2^, respectively), and 4.8% of cases had diabetes, compared to 2.6% of controls. Inflammatory markers at blood draw were not substantially different between cases and controls in each cohort, as previously reported.^6^ Pancreatic cancer cases from the PanGenEU study were older (mean 66.3 years old), and prevalence of current smoking and diabetes mellitus was also higher in that study (Table 1).

In the nested case-control study, immune cell proportions estimated from DNA methylation data did not vary by case-control status (Supplemental Figure 1). Furthermore, immune cell ratios for CD4/CD8, NLR, B-cell/lymphocyte, T-cell/lymphocyte, and monocyte/lymphocyte were not associated with risk of pancreatic cancer (Table 2). Associations were similar across cohorts, and among cases, the NLR remained stable as time from blood draw to diagnosis decreased (including blood draw ≤5 years prior to diagnosis). This analysis could not be conducted in the PanGenEU study as the DNA methylation was performed on granulocytes only.

**Table 2.** Odds ratios (OR) for immune cell ratio and pancreatic cancer risk in cohorts (nested case-control study).

|  | Age-adjusted Model |  | Multivariate-adjusted Model<sup>a</sup> |  |
| --- | --- | --- | --- | --- |
|  | Cases/Controls | OR (95% CI) | Case/Controls | OR (95% CI) |
| **CD4/CD8 Ratio** |  |  |  |  |
| Q1 (< 1.25) | 97 / 108 | ref. | 96 / 108 | ref. |
| Q2 (1.26 - 1.88) | 101 / 107 | 1.06(0.72, 1.56) | 100 / 107 | 1.07(0.72, 1.58) |
| Q3 (1.89 - 2.73) | 91 / 108 | 0.95(0.64, 1.41) | 90 / 106 | 0.97(0.65, 1.45) |
| Q4 (≥ 2.74) | 104 / 108 | 1.08(0.73, 1.58) | 104 / 107 | 1.10(0.74, 1.62) |
|  |  | p for trend = 0.84 |  | p for trend = 0.76 |
| **Neutrophil/Lymphocyte Ratio** |  |  |  |  |
| Q1 (< 1.29) | 97 / 108 | ref. | 97 / 108 | ref. |
| Q2 (1.30 - 1.69) | 82 / 107 | 0.85(0.57, 1.27) | 82 / 106 | 0.85(0.57, 1.27) |
| Q3 (1.70 - 2.26) | 108 / 108 | 1.11(0.76, 1.63) | 106 / 107 | 1.09(0.74, 1.61) |
| Q4 (≥ 2.27) | 106 / 108 | 1.09(0.75, 1.60) | 105 / 107 | 1.10(0.75, 1.61) |
|  |  | p for trend = 0.40 |  | p for trend = 0.41 |
| **B cell /Lymphocyte Ratio** |  |  |  |  |
| Q1 (< 0.10) | 96 / 108 | ref. | 96 / 106 | ref. |
| Q2 (0.11 - 0.13) | 84 / 107 | 0.89(0.60, 1.32) | 83 / 106 | 0.87(0.59, 1.30) |
| Q3 (0.14 - 0.17) | 100 / 108 | 1.05(0.71, 1.55) | 100 / 108 | 1.04(0.71, 1.54) |
| Q4 (≥ 0.18) | 113 / 108 | 1.19(0.81, 1.75) | 111 / 108 | 1.15(0.78, 1.71) |
|  |  | p for trend = 0.26 |  | p for trend = 0.34 |
| **T cell/Lymphocyte Ratio** |  |  |  |  |
| Q1 (< 0.58) | 103 / 108 | ref. | 102 / 108 | ref. |
| Q2 (0.59 - 0.64) | 95 / 107 | 0.94(0.64, 1.38) | 93 / 106 | 0.94(0.63, 1.39) |
| Q3 (0.65 - 0.69) | 101 / 108 | 0.99(0.68, 1.46) | 101 / 108 | 1.01(0.69, 1.50) |
| Q4 (≥ 0.70) | 94 / 108 | 0.93(0.63, 1.37) | 94 / 106 | 0.97(0.65, 1.44) |
|  |  | p for trend = 0.78 |  | p for trend = 0.97 |

<sup>a</sup> Adjusted for age, cohort, date of blood draw, and smoking.

**Figure 1.**
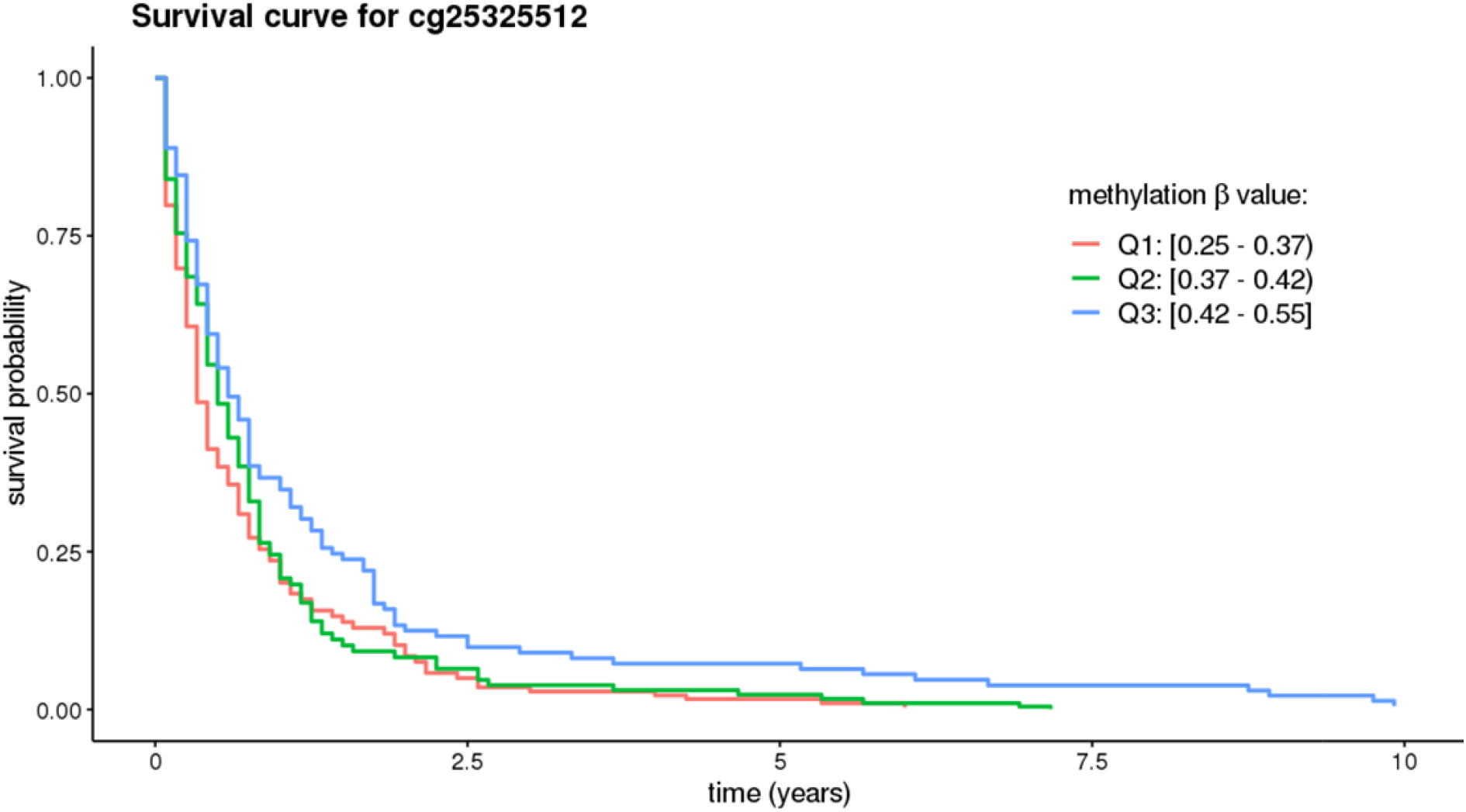
Survival curves among cases in the nested case-control study for the the CpG in PIM-1 promoter. Results for this CpG were consistent in the nested case-control study and PanGenEU. Curves are adjusted age, date of blood draw, time between blood draw and diagnosis, smoking, cohorts, and immune cell proportions.

50 CpG sites whose methylation extents were previously associated with inflammatory markers were examined in relation to pancreatic cancer risk in this dataset (Table 3). Of those, the methylation extents of 2 CpG sites (cg05304729 and cg06192883) were strongly associated with risk of pancreatic cancer overall in the nested-case control study (p<0.01 for trend across quartiles, without adjustment for multiple comparisons), and associations were consistently positive in at least 2 of the 3 cohorts (Table 3). For cg05304729, associations with pancreatic cancer were stronger when blood draw was closer to diagnosis (Table 3). Similar associations were noted for overweight or normal weight participants for both CpG sites. The positive trend for methylation extent of cg05304729 was replicated in the PanGenEU study; a significant test for trend was observed (p=0.01), with a 2-fold increase in risk in the highest quartile of DNA methylation (Table 3). However, no association was observed for methylation of cg06192883 and pancreatic cancer risk in the PanGenEU study (Table 3). Statistically significant results for the PanGenEU study are provided in Supplemental Table 2.

**Table 3.** Odds ratios for inflammatory-related CpGs and pancreatic cancer risk identified in the nested case-control study, stratified by study and time to diagnosis

| **cg05304729** |  |  | **cg06192883** |  |
| --- | --- | --- | --- | --- |
|  | Cases / Controls | Multivariate OR (95% CI)<sup>a</sup> | Cases / Controls | Multivariate OR (95% CI)<sup>a</sup> |
| **Among All Cohorts** |  |  |  |  |
| Q1 | 72 / 108 | ref. | 75 / 107 | ref. |
| Q2 | 93 / 107 | 1.37(0.90, 2.07) | 91 / 107 | 1.21(0.79, 1.83) |
| Q3 | 100 / 105 | 1.52(1.00, 2.31) | 84 / 106 | 1.12(0.73, 1.73) |
| Q4 | 125 / 108 | 1.92(1.27, 2.91) | 140 / 108 | 1.82(1.18, 2.78) |
|  |  | p for trend = 0.002 |  | p for trend = 0.008 |
| **Among Time to Diagnosis ≤ 5 Years** |  |  |  |  |
| Q1 | 6 / 108 | ref. | 6 / 107 | ref. |
| Q2 | 12 / 107 | 2.15(0.75, 6.16) | 11 / 107 | 1.83(0.63, 5.33) |
| Q3 | 12 / 105 | 2.14(0.74, 6.17) | 12 / 106 | 1.80(0.62, 5.24) |
| Q4 | 20 / 108 | 3.37(1.23, 9.18) | 21 / 108 | 2.57(0.92, 7.21) |
|  |  | p for trend = 0.02 |  | p for trend = 0.09 |
| **Among Time to diagnosis 5 - 10 Years** |  |  |  |  |
| Q1 | 11 / 108 | ref. | 15 / 107 | ref. |
| Q2 | 26 / 107 | 2.75(1.25, 6.07) | 23 / 107 | 1.68(0.79, 3.60) |
| Q3 | 18 / 105 | 2.27(0.97, 5.29) | 15 / 106 | 1.06(0.46, 2.45) |
| Q4 | 28 / 108 | 3.34(1.48, 7.54) | 30 / 108 | 1.87(0.85, 4.10) |
|  |  | p for trend = 0.01 |  | p for trend = 0.25 |
| **Among Time to diagnosis > 10 Years** |  |  |  |  |
| Q1 | 51 / 108 | ref. | 51 / 107 | ref. |
| Q2 | 53 / 107 | 1.11(0.68, 1.79) | 55 / 107 | 1.08(0.67, 1.74) |
| Q3 | 66 / 105 | 1.38(0.85, 2.21) | 56 / 106 | 1.13(0.69, 1.86) |
| Q4 | 76 / 108 | 1.64(1.02, 2.64) | 84 / 108 | 1.67(1.03, 2.72) |
|  |  | p for trend = 0.03 |  | p for trend = 0.03 |
| **Among NHS<sup>b</sup>** |  |  |  |  |
| Q1 | 44 / 49 | ref. | 35 / 49 | ref. |
| Q2 | 39 / 48 | 0.88(0.48, 1.63) | 32 / 48 | 0.82(0.43, 1.57) |
| Q3 | 45 / 48 | 1.07(0.58, 1.98) | 42 / 48 | 1.09(0.57, 2.06) |
| Q4 | 47 / 49 | 1.04(0.56, 1.96) | 66 / 49 | 1.53(0.80, 2.95) |
|  |  | p for trend = 0.75 |  | p for trend = 0.11 |
| **Among HPFS<sup>b</sup>** |  |  |  |  |
| Q1 | 20 / 38 | ref. | 31 / 37 | ref. |
| Q2 | 32 / 37 | 1.79(0.86, 3.73) | 44 / 36 | 1.49(0.74, 3.01) |
| Q3 | 35 / 36 | 1.95(0.92, 4.11) | 26 / 37 | 0.82(0.38, 1.76) |
| Q4 | 57 / 37 | 3.44(1.67, 7.12) | 43 / 38 | 1.33(0.64, 2.77) |
|  |  | p for trend <0.001 |  | p for trend = 0.85 |
| **Among PHS<sup>b</sup>** |  |  |  |  |
| Q1 | 9 / 22 | ref. | 7 / 22 | ref. |
| Q2 | 22 / 21 | 2.58(0.94, 7.11) | 20 / 21 | 3.49(1.16, 10.55) |
| Q3 | 19 / 21 | 2.49(0.89, 7.01) | 24 / 21 | 5.07(1.66, 15.47) |
| Q4 | 21 / 22 | 2.62(0.93, 7.36) | 20 / 22 | 3.65(1.20, 11.10) |
|  |  | p for trend = 0.12 |  | p for trend = 0.03 |
| **Replication in PanGenEU<sup>b,c</sup>** |  |  |  |  |
| Q1 | 50 / 71 | ref. | 72 / 71 | ref. |
| Q2 | 77 / 71 | 1.49(0.9, 2.49) | 60 / 71 | 0.82(0.49, 1.35) |
| Q3 | 86 / 71 | 1.48(0.88, 2.48) | 94 / 71 | 1.23(0.77, 1.98) |
| Q4 | 126 / 72 | 2.08(1.22, 3.57) | 112 / 72 | 1.33(0.80, 2.21) |
|  |  | p for trend = 0.01 |  | p for trend = 0.11 |

<sup>a</sup>Adjusted for age, date of blood draw, smoking and cell proportions, and cohorts for combined analyses.
<sup>b</sup>Used study-specified quartiles for methylation level; <sup>c</sup> PanGenEU model adjustments include age, sex, smoking and cell proportions.

We also examined whether the immune cell ratios were associated with survival time among the cases in the nested case-control study (Table 4). Overall, the immune ratio measures were not associated with survival time, and associations were similar when stratifying on time between blood collection and date of diagnosis. Among the 50 CpGs tested, methylation level of 6 CpGs were statistically significant associated with survival at p≤0.05 (cg00159243, cg03957124, cg12785694, cg1818703, cg25325512, cg26804423; Table 4). Methylation level at two of these CpGs (cg00159243, cg25325512) was significantly associated with risk in PanGenEU (Supplemental Table 2), and methylation of cg25325512 was also associated with survival in PanGenEU (Q2 vs Q1: HR= 0.71, 95% CI 0.52-0.96; Q3 vs Q1: HR 0.72, 95% CI 0.51-1.00, p-trend 0.057). Survival curves for methylation levels at this CpG in the cases from the cohort study are presented in Figure 1. The ICC for cg25325512 was 0.86 in our pilot study (over a 1-year period), suggesting that methylation at this probe does not vary much over time, and thus providing a valid proxy for levels closer to diagnosis. Of note, cg25325512 is located in the PIM-1 gene which has previously been associated with survival of pancreatic cancer.

**Table 4.**
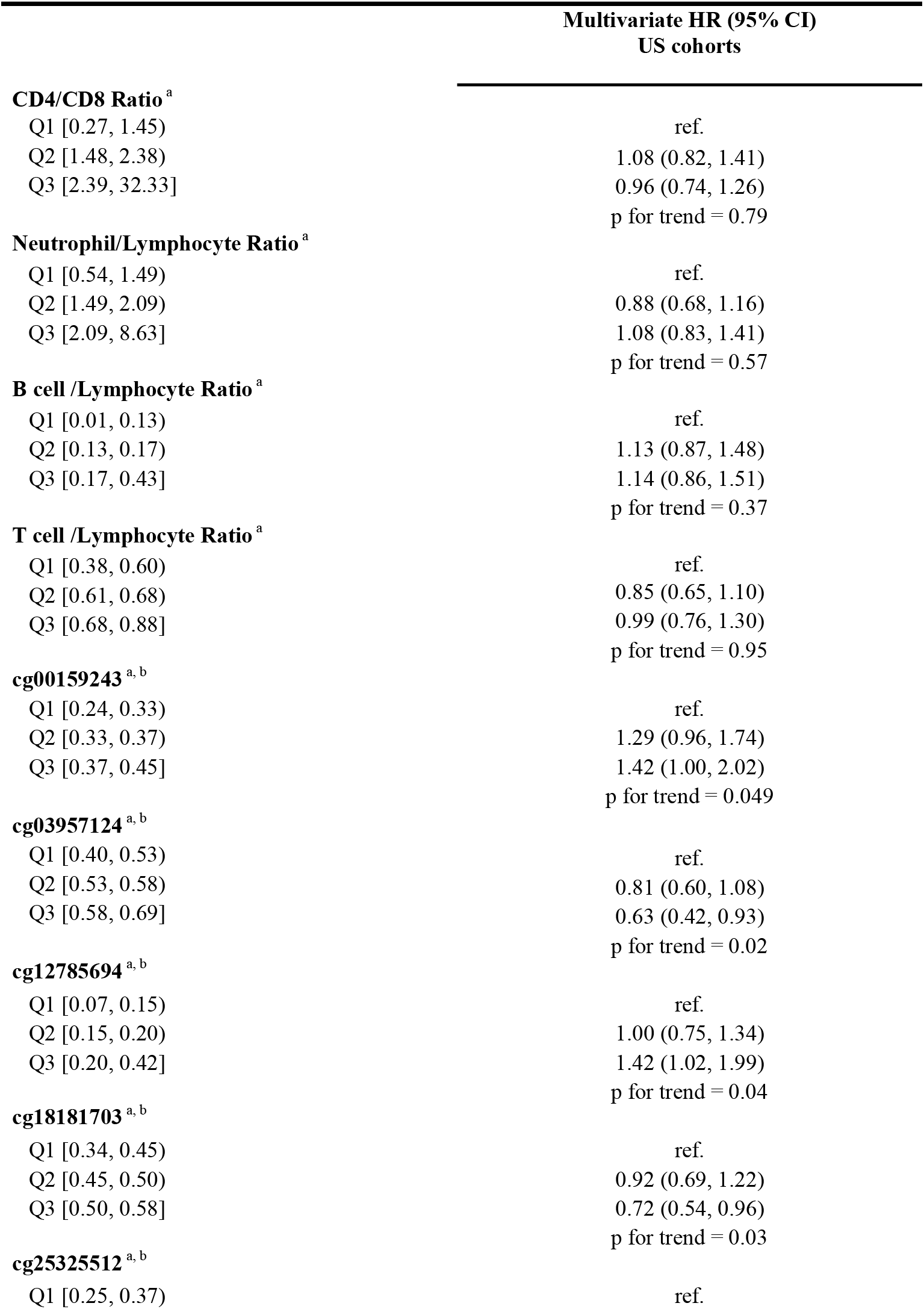
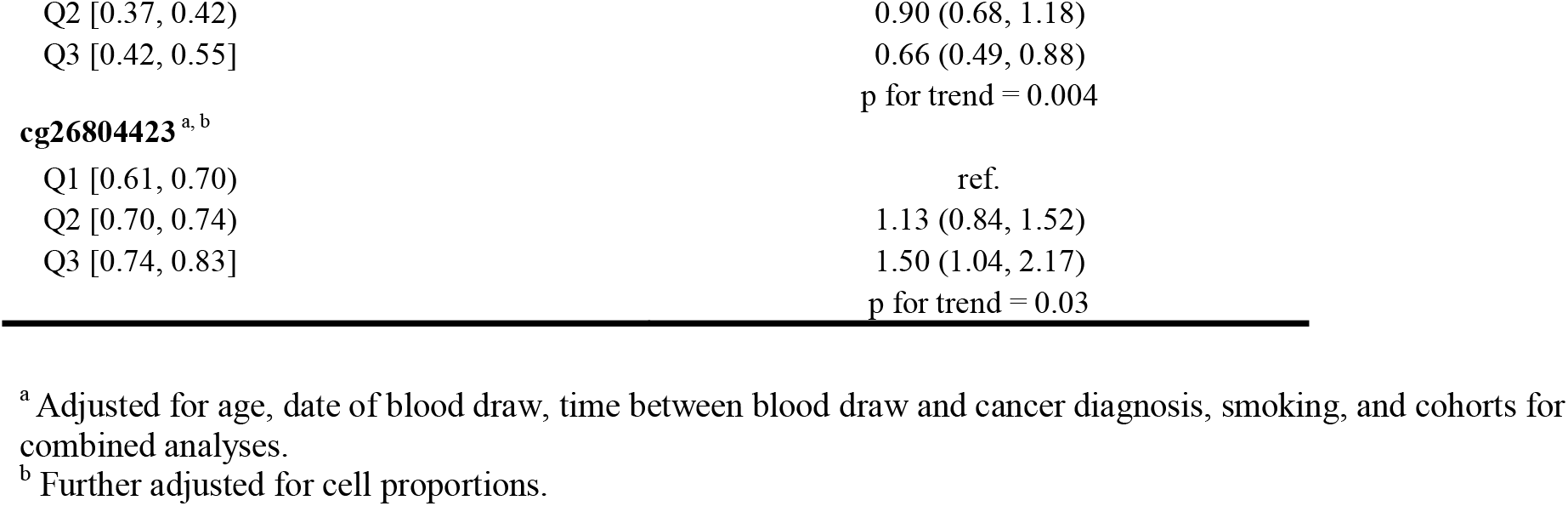
Association between immune cell counts ratio, inflammatory-related CpGs and pancreatic cancer survival time among cases from cohorts only (n = 342)

## Discussion

To our knowledge, this is the first study to examine associations between CpG methylation of inflammation markers, methylation derived immune cell composition, and risk of pancreatic cancer using pre-diagnostic blood samples. One of the goals of this study was to measure immune cell proportions in blood samples using established DNA methylation markers of immune cell types as flow cytometry could not be conducted on archived frozen bloods. While we did not find any associations for ratios of immune cell proportions, we did identify and replicate an association with the DNA methylation level of a CpG previously associated with inflammation.

Epigenetic-wide association studies (EWAS) using Illumina arrays to identify methylation at CpG sites associated with inflammatory blood markers have been carried out in two large studies.^15, 16^ We selected 50 CpG sites that had met criteria for inclusion in this analysis and identified two (cg05304729 and cg06192883) that were statistically significantly associated with pancreatic cancer risk in the nested case-control study overall. Furthermore, for cg05304729, the associations were stronger as the collection of bloods got closer to date of diagnosis, suggesting the inflammation increased closer to diagnosis, perhaps due to subclinical changes. The fact that the association was present 10 years prior to cancer diagnosis (Q4 vs Q1 OR =1.64, 95% CI = 1.02, 2.64; Table 3) suggests that the methylation level at that site is related to risk, rather than being sole consequence of the cancer. In a separate replication analysis using a case-control study with blood collected at pancreatic cancer diagnosis (PanGenEU), we observed similar associations for cg05304729. However, we did not observe an association for cg06192883 in PanGenEU, suggesting this finding may have been a chance finding.

Previous studies have reported strong associations between methylation at cg05304729 and levels of three different inflammation markers measured in blood (CXCL9^15^, CXCL11^15^ and TNFRSF6B^18^). In our study, methylation at cg05304729 was not correlated with CRP, TNFαR2, or IL-6 (Supplemental Table 1). DNA methylation at both CpG sites have also been associated with BMI,^18^ out of the 102 CpG sites tested in Myte et al., the two CpG sites identified in our current study were among the three most statistically significant associations with BMI in the prior study (p-values =0.0001). In addition, cg05304729 was identified as 1 of 20 probes associated with BMI in a separate EWAS study (FDR q = 0.015)^19^ and cg06192883 was identified in another EWAS study on BMI.^20^ Given the known role of BMI in pancreatic cancer risk, the DNA methylation sites identified in this study may provide insight into the underlying biological pathways involved. Cg05304729 is located 200-1500 bases upstream of the transcriptional start site (Illumina annotation: TSS1500) for the myeloid nuclear differentiation antigen (*MNDA*) gene; expression of this gene has been previously associated with lymphoma, especially marginal zone derived lymphomas.^21^ This gene may also be involved in cell-specific response to interferons.^22^ More research will be necessary to understand the role of these pathways in pancreatic cancer.

Conducting a survival analysis, we identified methylation level for two CpG sites (cg00159243, cg25325512) that were significantly associated with survival of pancreatic cancer in the nested case-control study (p≤0.05), and significantly associated with risk in PanGenEU (p<0.05). However, only the extent of methylation of cg25325512 was also associated with survival in PanGenEU (p=0.057). Cg25325512 is located on gene PIM1, a well-established oncogene^23^ that has been widely targeted for anticancer drug discovery.^24^ Some studies have shown that high PIM-1 expression in pancreatic tumor tissue is associated with worse survival and, in a recent study, *plasma* PIM-1 level was associated with pancreatic cancer survival (HR = 1.87, 95% CI = 1.04-3.35) and risk (p<0.0001).^25^ Given the implication of this finding, we went back to examine whether the association with risk existed in the nested case-control study (i.e., including controls); although the trend test was not significant, the highest quartile was borderline significant (HR = 0.68, 95% CI = 0.44 - 1.05, compared to the lowest) overall, and significant when blood was collected 10 years prior to diagnosis (HR = 0.55, 95% CI = 0.34 - 0.91, top to bottom quartile comparison). This finding is particularly interesting as it suggests DNA methylation at this site occurred many years prior to diagnosis and thus is not likely to be caused by the tumor development.

Our study strengths include use of pre-diagnostic bloods and a large number of incident pancreatic cancer cases. Pre-diagnostic bloods are critical to determine whether methylation states at different CpG sites were present prior to diagnosis, rather than identifying changes that might have occurred as a result of the cancer. By ruling out reverse causation, we could begin to identify pathways that play a role in the etiology of the disease but also identify early diagnosis markers. Being able to examine associations in a separate case-control study (PanGenEU) was an additional strength to this analysis as it provided an opportunity to evaluate the robustness of our findings in a completely different population, providing strong evidence of reproducibility. Other strengths of this study included adjustment for potential confounders, including age, race, smoking, BMI, and diabetes. Moreover, our data processing steps and random assignment of samples on plates removed potential technical biases.

Study limitations include our reliance on established DNA methylation markers for immune cell types, which are primarily limited to the main immune cell types. Subsets of immune cells that are more difficult to identify and may play a role in cancer, such as regulatory T-cells, could be associated with cancer risk, but were not available for this analysis.

This is the first prospective study examining the associations between immune cell proportions and risk of pancreatic cancer. While we did not observe associations with risk for several main know indicators of immune status previously associated with survival, such as NLR, we identified two CpGs that have been strongly associated with inflammation and BMI in prior studies. More research on *MNDA* and *PIM-1* genes may reveal new area of research for pancreatic cancer risk, given that these genes have been previously implicated in other cancers, and PIM-1 expression has previously been associated with lower pancreatic cancer survival. Further research based on our findings may lead to identification of novel proteins that are differentially expressed prior to cancer diagnosis that could be tested in blood for early detection or for the identification of individuals at higher risk (without the need for DNA methylation measurements). Alternatively, our findings could lead to identification of pathways that may be targetable for treatment.

## Data Availability

All data from this study have been deposited in dbGAP and will be available on January 3, 2020 [“DNA Methylation Markers and Pancreatic Cancer Risk in 3 Cohort Studies (NHS, PHS, HPFS)” phs001917.v1.p1].

https://www.ncbi.nlm.nih.gov/projects/gapprev/gap/cgi-bin/study.cgi?study_id=phs001917.v1.p1

## Data Availability Statement

All data from this study have been deposited in dbGAP and will be available on January 3, 2020 [“DNA Methylation Markers and Pancreatic Cancer Risk in 3 Cohort Studies (NHS, PHS, HPFS)” phs001917.v1.p1]. https://www.ncbi.nlm.nih.gov/projects/gapprev/gap/cgi-bin/study.cgi?study_id=phs001917.v1.p1

## Acknowledgment

**PanGenEU centers and investigators**

<u>Spanish National Cancer Research Centre (CNIO), Madrid, Spain</u>: Núria Malats^1^, Francisco X Real^1^, Evangelina López de Maturana, Paulina Gómez-Rubio, Esther Molina-Montes, Lola Alonso, Mirari Márquez, Roger Milne, Ana Alfaro, Tania Lobato, Lidia Estudillo.

<u>Verona University, Italy</u>: Rita Lawlor^1^, Aldo Scarpa, Stefania Beghelli.

<u>National Cancer Registry Ireland, Cork, Ireland</u>: Linda Sharp^1^, Damian O’Driscoll.

<u>Hospital Madrid-Norte-Sanchinarro, Madrid, Spain</u>: Manuel Hidalgo^1^, Jesús Rodríguez Pascual. <u>Hospital Ramon y Cajal, Madrid, Spain</u>: Alfredo Carrato^1^, Carmen Guillén-Ponce, Mercedes Rodríguez-Garrote, Federico Longo-Muñoz, Reyes Ferreiro, Vanessa Pachón, M Ángeles Vaz.

<u>Hospital del Mar, Barcelona, Spain</u>: Lucas Ilzarbe^1^, Cristina Álvarez-Urturi, Xavier Bessa, Felipe Bory, Lucía Márquez Mosquera, Ignasi Poves Prim, Fernando Burdío, Luis Grande, Mar Iglesias, Javier Gimeno.

<u>Hospital Vall d’Hebron, Barcelona, Spain</u>: Xavier Molero^1^, Luisa Guarnert, Joaquin Balcells. <u>Technical University of Munich, Germany</u>: Christoph Michalski^1^, Jörg Kleeff, Bo Kong.

<u>Karolinska Institute, Stockholm, Sweden</u>: Matthias Löhr^1^, Jiaqui Huang, Weimin Ye, Jingru Yu.

<u>Hospital 12 de Octubre, Madrid, Spain</u>: José Perea^1^, Pablo Peláez.

<u>Hospital de la Santa Creu i Sant Pau, Barcelona, Spain</u>: Antoni Farré^1^, Josefina Mora, Marta Martín, Vicenç Artigas, Carlos Guarner Argente, Francesc J Sancho, Mar Concepción, Teresa Ramón y Cajal.

<u>The Royal Liverpool University Hospital, UK</u>: William Greenhalf^1^, Eithne Costello. <u>Queen’s University Belfast, UK</u>: Michael O’Rorke^1^, Liam Murrayt, Marie Cantwell.

<u>Laboratorio de Genética Molecular, Hospital General Universitario de Elche, Spain</u>: Víctor M Barberá^1^, Javier Gallego.

<u>Instituto Universitario de Oncología del Principado de Asturias, Oviedo, Spain</u>: Adonina Tardón^1^, Luis Barneo.

<u>Hospital Clínico Universitario de Santiago de Compostela, Spain</u>: Enrique Domínguez Muñoz^1^, Antonio Lozano, Maria Luaces.

<u>Hospital Clínico Universitario de Salamanca, Spain</u>: Luís Muñoz-Bellvís^1^, J.M. Sayagués Manzano,

M.L. Gutíerrrez Troncoso, A. Orfao de Matos.

<u>University of Marburg, Department of Gastroenterology, Phillips University of Marburg, Germany</u>: Thomas Gress^1^, Malte Buchholz, Albrecht Neesse.

<u>Queen Mary University of London, UK</u>: Tatjana Crnogorac-Jurcevic^1^, Hemant M Kocher, Satyajit Bhattacharya, Ajit T Abraham, Darren Ennis, Thomas Dowe, Tomasz Radon

<u>Scientific advisors of the PanGenEU Study</u>: Debra T Silverman (NCI, USA) and Douglas Easton (U. of Cambridge, UK)

## Footnotes

1 Principal Investigator in each center

